# Analysing recovery from pandemics by Learning Theory: the case of CoVid-19

**DOI:** 10.1101/2020.04.10.20060319

**Authors:** Romney B Duffey, Enrico Zio

## Abstract

We present a method for predicting the recovery time from infectious diseases outbreaks such as the recent CoVid-19 virus. The approach is based on the theory of learning from errors, specifically adapted to the control of the virus spread by reducing infection rates using countermeasures such as medical treatment, isolation, social distancing etc. When these are effective, the infection rate, after reaching a peak, declines following a given recovery rate curve. We use presently available data from China, South Korea and others to make actual predictions of the time needed for securing minimum infection rates in the future.

## 1. Introduction and Background

We develop a new way to predict the recovery rate of infections following a pandemic outbreak, using the basic postulates of learning theory. This theory has been previously applied to outcome, accident and event data from multiple socio-technological systems, like transportation, medicine, military, power grids, aviation, mining, and manufacturing. Learning theory simply postulates that humans learn from experience in correcting their mistakes and errors (sometimes even just by trial and error), as they gain knowledge on the problem and skill for addressing it. The theory is consistent with the models and data in cognitive psychology of how humans behave and the brain operates (Ohlsson, 1996; Fiondella and Duffey, 2015; Anderson, 1990; Duffey, 2017b). The importance of this theory stands in that human errors and incorrect decisions are the dominant contributors to accidents, crashes, system failures, errors, and operational incidents.

The theory is based on the fact that human learning demonstrably reduces error rates (Ohlsson, 1996): wisdom is gained after an accident. Evidence on this relates to even highly hazardous industries like the nuclear one. A good example is that the safe operation of nuclear power plants has been, and continuous to be, improved from lessons learned from nuclear accidents and incidents. These accidents and incidents, in addition to highlighting the role of human errors in their occurrence and progression, have helped identifying various critical technical elements and contributed to the safer operation of nuclear power plants. Similarly, the observation is applicable to outbreaks of infectious diseases.

Improving health systems following epidemic outbreaks and enhancing reliability and safety measures following nuclear power plant accidents have to be handled with objective data and accurate calculations. However, whereas nuclear power plant operation has done this by a world-accepted, high standard of procedures, the “protection system” against pandemics is not there yet. The key is how to design, evaluate and implement the procedures, for reaching a high standard.

Learning theory has been successfully applied to quantify error and learning rates in technical systems, casualties in large land battles, everyday accident and event data, and to human, software and hardware reliability (Duffey and Saull, 2008; Duffey & Ha, 2010; Fiondella & Duffey, 2015; Duffey, 2017a). The novel feature is to replace calendar time or test interval, which has always been used before, with a measure for the accumulated experience and/or risk exposure, thus defining rate trends and quantifying effectiveness of responses to errors and accidents, and allowing totally different systems to be directly intercompared. Additionally, the trend is governed by two parameters that are physically based: the learning rate constant and the minimum achievable error rate. This is in contrast with statistical analysis, where fitting to learning data is typically done on three empirical parameters (Bush and Mosteller, 1955), and with the inverse “power laws” extensively fitted in cognitive psychology data (e.g. Anderson, 1990 and the references therein).

The theory shows the Learning Hypothesis that humans learn from their mistakes and reduce outcomes in such a way that the rate of decrease of the outcome (in the present case of interest, the infection rate, R) with the rate of accumulated experience, ε, (in the present case the advancement in the knowledge of the virus, the contagion spreading dynamics, the effects of the countermeasures) is proportional to the rate R itself. Thus, very simply, the differential equation that describes the accident and outcome data with learning or forgetting describes the proportionality between the rate of change of the learning *rate*, R, and the learning rate itself (Duffey and Saull, 2008; Duffey, 2017b):

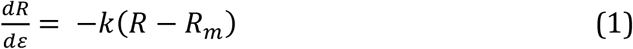

where ε is the measure of the risk exposure, learning opportunity or experience/knowledge gained; k is the learning rate (positive for a learning/improving situation and negative for no learning/improving, e.g. because of no effectiveness countermeasures) and R_m_ is the lowest or minimum achievable error rate, which is never zero as the process of error-making and cognitive rule revision always continues. Physically, k is related to the non-detection or error rate in unconscious memory scanning for recall and recognition, manifesting itself in the conscious external actions, decisions and judgments. The error rate solution obtained from integration of this Minimum Error Rate equation (MERE) is:

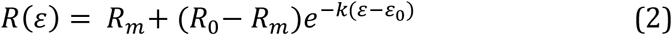

where, *R*_0_is the initial rate at the beginning or start of the problem when the level of experience/knowledge on it is ε= ε_0_. Different data sets are characterized by different values of the learning constant.

It is clear in Equation (2) that for practical applications a suitable measure of the experience/knowledge or risk exposure accumulated with respect to the initial one, (ε-ε_0_), is needed and that the original or starting one, ε_0_, is a suitable arbitrary or convenient reference dependent on the problem at hand. In other words, the measure for the accumulated experience/knowledge or risk exposure is technology/system specific. Then, for data inter-comparisons, it is useful to render non-dimensional the quantities of interest, which results in the Universal Learning Curve (ULC) for the non-dimensional error rate E*= (R(ε)-R_m_)/(R_0_-R_m_) as a function of the non-dimensional experience/knowledge or risk exposure ε* = ε/ ε_T_, where ε_T_ is the maximum accumulated experience or risk exposure thanks to which the error is recovered (the problem is considered under control), its rate having reached the lowest or minimum achievable value R_m_. From (2),

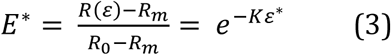

with K∼ 3 is the fitted learning rate “universal constant”. This expression has already been shown to represent the learning trends for outcome rate data from industrial, surgical, transportation, mining, manufacturing, chemical, maintenance, software and a multitude of other systems (Duffey and Saull, 2008; Duffey and Ha, 2013: Fiondella and Duffey, 2015). For skill acquisition tasks in cognitive psychological testing, this same trend exists and is called the Universal Law of Practice (ULP).

## 2. “Normal” Infectious disease risk

Illnesses are still around in the world, many of them deadly. In the past, there have been pandemics^1^ killing many millions of people, like the “Black Death” or Bubonic Plague disease of the Middle Ages, and the influenza epidemic in 1918. In addition to these sudden attacks, other equally deadly pestilences have been and are still around for centuries - yellow fever, cholera, small pox, typhus, measles, malaria As modern medical practice eliminated or reduced these hazards using better procedures and new vaccines, other exotic variants and viruses have recently emerged, like SARS, HIV, Ebola and CoViD19, infecting and endangering the ever-increasing and interconnected world population. As we evolve and learn, so do the things that like to kill us, but they usually kill relatively few people compared to, say, automobile accidents or the yearly seasonal influenza.

To determine risk from these instances, we can and must turn to data. As a fine example, we have the official data from the World Health Organization. The WHO gave the death rates for “all causes” and for infectious (like cholera) and parasitic (like malaria) diseases for some 194 countries in the mid 1990s. The data cover the full global spectrum, from developed to developing nations, from vast urban conglomerates with very crowded living conditions to scattered rural communities, from jungles to deserts, and all Continents. The data cover and include the effects of modern epidemics, and of course local wars and regional conflicts.

In Figure 1, the data are plotted *not* against the usual arbitrary calendar year but- as we now know we should-against the risk exposure measure, in this case the population size. This number is the direct indicator of how many people are at risk, and the country-by-country populations come from the World Bank Indicators. The data are plotted in the Figure with the lozenges being the overall death rate data and the squares representing the death rate data due to infections.

**Figure 1.**
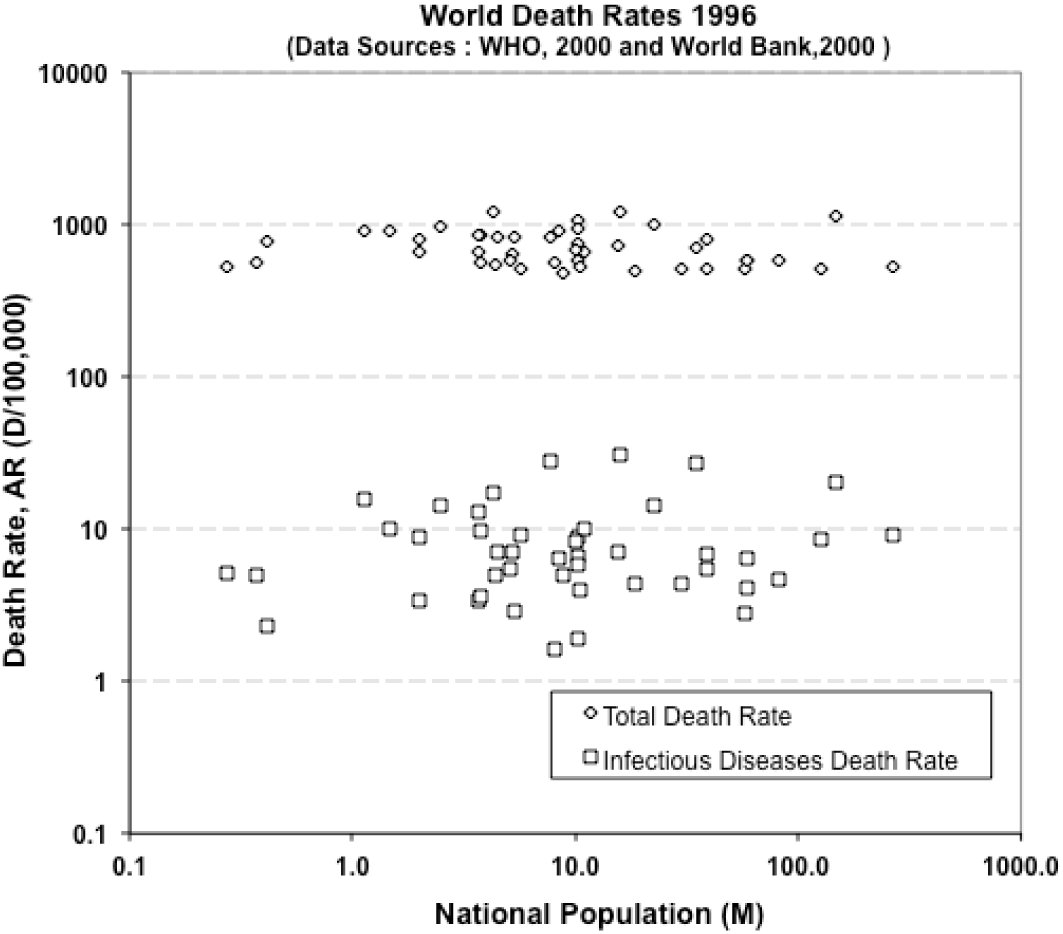
The rate of deaths from all causes (lozenges) and from infectious diseases (squares) for 194 countries (data extracted from the WHO and World Bank).

From these data, we can know the risk of death from any health cause: it is about 1000 deaths for every 100,000 people, or one in a hundred, and does not depend much on where you live. To verify this overall number “locally”, we can analyze the data for New York, as given in the graph “The Conquest of Pestilence in New York City” from 1800 onwards, published by the Board of Health and the Health Department. This is a typical modern city that had grown in population from 120,000 to about 8 million people, and includes characteristics of immigration, high-density living, mass transportation, high-rise apartments, modern health care, national and international trade, and a large flow of inbound-outbound travel, in other words, globalization. The biggest improvement in health has come from introducing effective hygiene and anti–infection measures, and from improved health prevention and treatment (not from wonder drugs): we have learned how to treat sick people, cure problems and reduce the spread of bad diseases. It is an expensive investment and it is hard work that requires devoted and trained professionals. As a result, after curing and containing many pestilences during the 19^th^ century, the average death rate in New York over the last hundred years has fallen to 10 to 11 per 1000 people, or almost exactly the same one-in-a-hundred rate as the world rate. So modern cities behave pretty much like whole countries, as far as average or overall death rates are concerned.

Infectious and parasitic diseases are responsible for 5 to 15 deaths in 100,000 people, so average about one in ten thousand or 1 to 5% of all deaths worldwide, the other 95 to 99% or so being deaths from “normal” causes. So the “normal” death risk is still about twenty to a hundred times of what it might be if a new pestilence emerges, spreads and takes hold without effective countermeasures.

Another way to view this risk contribution is to say that the chance of death might be increased by a maximum of about 5% if a new global pandemic infection occurs where it has not been prevalent before. This is always the fear, that in today’s highly interconnected, high-speed, global world a possible rapid spread of new or variant diseases can occur.

And that is exactly what recently happened, as the 2019-2020 coronavirus (labeled CoVid- 19 or SARS-CoV2) rapidly spread across the four corners of the Globe. There was extensive reporting of nearly every new case occurring and great worldwide information available (e.g. at Johns Hopkins University website coronavirus.jhu.edu/map). By late March 2020, the world had over 1,000,000 cases (and still growing at the time of writing) and many thousands of deaths, with the infection having spread quickly across borders, imported from nation-to-nation mainly via travellers, visitors and tourists, and spread internally from just social and day-to-day human contact.

## 3. Infection rate reduction and recovery

Looking at the available numbers, the *individual* infection risk today is comparatively negligible, with a few hundred thousand cases in a world population of several billion. It is an individual random infection probability, p(I), of about one in twenty thousand. This risk is many times less than the chance of being hospitalized with influenza, let alone catching it, reflecting the existing fraction of world deaths due to infections.

But the high speed at which this virus has spread makes it legitimate to feel worried and unease, and correspondingly, legitimate questions arise:

How does this novel pandemic compare to the “normal” or accepted risk of infectious death?

What is the “worst case” scenario?

Should we panic and shun other people who may be carrying what might kill us? How long will it take to recover?

This calls for the need to try to objectively evaluate the risk, based on the current experience knowledge.

We normally can treat the spread of disease as a “diffusion” or “multiple contact” process, where it steadily expands outward from some central source or origin; or as a highly mobile source that is potentially spread everywhere due to rapid multiple global personal and social interactions. The excellent US Centers for Disease Control simply states the obvious:

“Risk depends on characteristics of the virus, including how well it spreads between people”

(Source: www.cdc.gov/coronavirus/2019-ncov/summary)

The data from China on CoVid-19 infections suggest that there is, or was, a 50%-90% chance of the initial infections spreading between cities, depending on location and size (Du et al, 2020). Using simple doubling rates, the news media carried projections that a “worst case” in the USA could infect over 200 million (i.e. most people in the USA) and cause nearly 2 million or so deaths, and experts were hard at work estimating global, country and age-dependent risks, including of death.

We certainly need to estimate or know the risk of infection. As a simple guess and knowing nothing else, let us assume infections are randomly transmitted anywhere and everywhere from person to person, the spread is instantaneous and guaranteed if a source exists and the probability of being (successfully!) infected is also random and equally possible. This is really a “worst case” scenario or “model”, as obviously not everyone is exposed to everyone and not everyone is equally vulnerable. The worst case scenario is, then, that there is no preventative measures, no immunity and no vaccine, and for whatever reason, the source is not quarantined or isolated, and any such infected person or mobile source can and does transmit the virus (or disease) *randomly* to others somewhere in the world.

Independent of the transmission mechanism, given contact, the probability of cross-infection, then, depends solely on the total numbers of the possibly *equally risk- or infection-exposed* recipient population, and the probability of infection is also random. In this model, anyone can get it by interacting with someone that has it^2^.

To help see this more clearly, the black balls (the “unknowns”) emerging from the Jar of Life is one way to view what might happen based on what we have already seen or been exposed to. Here the one black ball (n=1) is an “known unknown” infected person or *infection opportunity* among those ten (m=10) non-infected (“known knowns”) white balls (m=10), so a chance of 10% or one over ten. The probability of interest is, then, of infection for more people exposed and infected, or N “unknown unknowns”, and some not exposed or not successfully infected, M “unknown knowns”), out of a total of all exposed people, N+M.

These numbers, N and M, vary by city, country, cruise ship passengers, soccer matches or rock concert arena, and can systematically vary up to the total of about six billions or so in the global world. We can also think of it as our possible exposure experience. The formula for the probability p of the event of an individual I becoming infected takes the form,

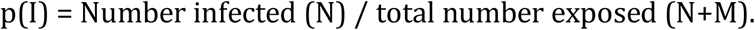

If there is a gathering limit of ten or so people, as in the case of the white and black balls of the Jar of Life shown in the picture, the observed chance of being infected, p(I), is assumed to be about one infection (black ball) in some ten non-infections (white balls), or 10%. This we know. But the sample is limited to 10 and, so, we could also be exposed to one in a hundred, or one in a thousand, or more: how many more infection cases would we, then, expect out there hidden in the Jar?

The method used to estimate this probability is random sampling based on the so-called hypergeometric formula^3^. For possible gathering numbers of 10,100, 300 and 1000 with one infection source known or observed, the chance of observing or finding, say, 100,000 new cases of infection, p(I) in a bigger group rises to a maximum or peak of about 37 % (about one in three)^4^.

The estimate that comes out in this “worst case scenario” seemingly agrees with the estimates publicly available on the internet and social media of the awful possibility (risk) of perhaps even a 40-70% chance of being at risk of infection, if nothing is done to prevent it or reduce it^5^. That is a significant and very high risk of infection, and has been used to justify quarantining, limiting social gatherings and extensive travel restrictions.

This inevitably brings fear to the individuals of the global population and the only way to address fear is by using scientific knowledge and data to inform any theory behind estimates and predictions.

During the early onset of the CoVid-19 pandemic, many gloomy scenarios were made and analyzed but they generally assumed no effective countermeasures to the spread of infections. The infection numbers grew quickly at first, before countermeasures such as isolation, distancing, restrictions and curfews were implemented to reduce infection rates and “flatten the curve” of numbers versus time. Sad to say, deaths (distressing as they are) are also NOT the right measure- infections are the measure for the spread and control of infectious diseases. A logical question is whether the infection or death data show any signs that we are learning how to reduce risk?

Just like for any accident- the number killed or dying is highly variable depending on who and how many risk-exposed happens to be there, so it is random. In this viral case, the number of deaths also just depends on too many uncontrolled variables and factors (age, pre-existing health conditions, health care system, propensity etc. …) so the average death percentage per infection also varies in magnitude, location and time (as the data clearly show). The correct measure is infection numbers and rates (not the number of deaths), say the number per day. The infections numbers also depend on which country/region they refer to and at what infectious stage (early onset, spread extent, countermeasures employed … etc.). We already know that if uncontrolled, the increase in infections will rise exponentially, as the rate of infections is proportional to the number infected.

As usual, the most representative case can be based on actual infection data from where containment of contagion has already been successfully applied, namely in China where it originated. Using data reported by the John Hopkins Center for System Science and Engineering, by March, 2020, there were, n= 81,000 infections and some 3200 deaths in China with a falling to near-zero rate. So with a national population of N+M =1,400,000,000, the overall probable risk of infection is about 0.00006, or one in 17,000, (or about six in 100,000)^6^. Locally, in some cities/regions it is ten times higher, but although comparable to infectious death rates, the overall CoVid-19 death rate in China was on average one in nearly half a million people, much lower than deaths from other infectious diseases. That is a significant reduction in the risk of infection: countermeasures have worked.

So far, we have these scenarios:

The scary or “Worst” with no measures: at least one in three people infected. The real or “Best” control and mitigation: only one in 17000 people infected. But remember the famous Bayes Theorem:

*The probable Future is the Past modified by the Likelihood*.

So the possible probability reduction, or Likelihood, must be considered in a future estimate based on projections using past data. This simple-minded upper and lower limit comparison suggests the individual Likelihood is about 0.0002, or very low. Societal countermeasures reduced the risk by a factor of 500 to 5000 over the inevitable “worst case” random spread of infection or survival-of–only- the-fittest scenarios.

## 4. Recovery timescales using Learning Theory

The approach based on Learning Theory illustrated in this Section uses the fact that humans learn how to control the CoVid-19 outbreak spread and reduce infection rates using countermeasures (treatments, isolation, “social distancing” etc). IF these are effective the rate, therefore, must reach a peak and, then, decline.

To look at pandemic recovery, we really need to look at the rate of infections, NOT just deaths since these depend on too many social and personal health factors as already stated.

These include propensity, age profile, medical system effectiveness, treatment options early detection, etc. The risk of an infectious disease is not controlled unless the infection rate slows down, so-called ‘flattening the curve”. Once the rate peaks the rate, then, should decrease due to successful countermeasures (whatever they are).

So, the next question is: what happens next and how do we know how effective the control measures are, and how long before they can be relaxed or maintained?

In China, the actual infection rates (increase per day) rose to a peak of nearly 4000 a day in about 17 to 20 days, and, then, fell away steadily to about 50 or so a day by another 30 days. As a further consideration for a different country, S Korea had a different peak rate of about 1000 per day in ten days, falling to a low rate in another 10 or so days.

In Italy, after some delay in implementing countermeasures, the infection rate seemingly peaked at about 6000 per day in about 30 or so days, as can be seen in the graph of Figure 3. As of writing, (early April) this rate has decreased to nearly 4000 per day (see graph in the Figure).

**Figure 2.**
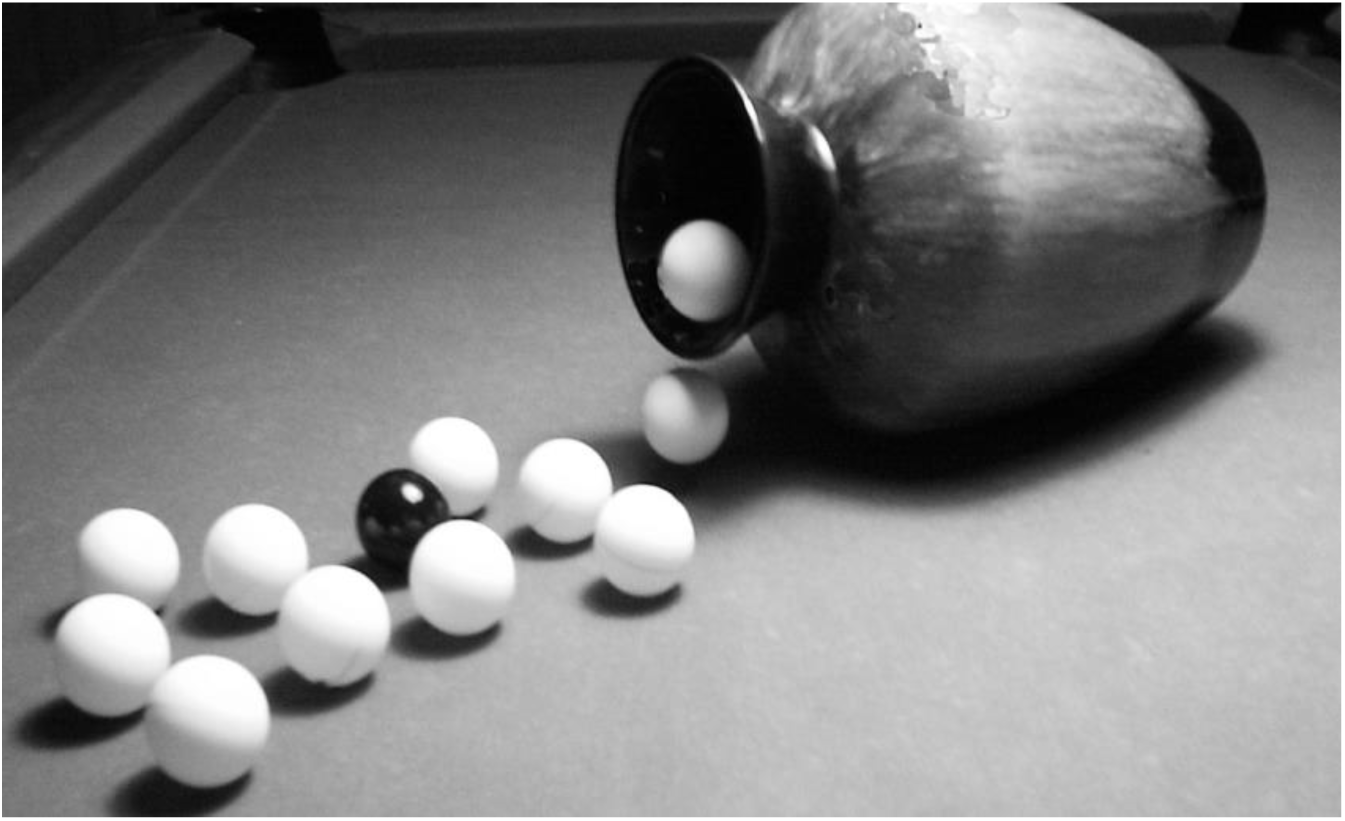
Randomness at work: we have already seen or heard about at least one infection (black ball) among our ten friends, colleagues, or contacts (white balls); so the obvious questions are how many more infections (black balls) are hidden in the jar, i.e. possible in the future, and what is the risk of drawing a black ball, i.e. of infection?

**Figure 3.**
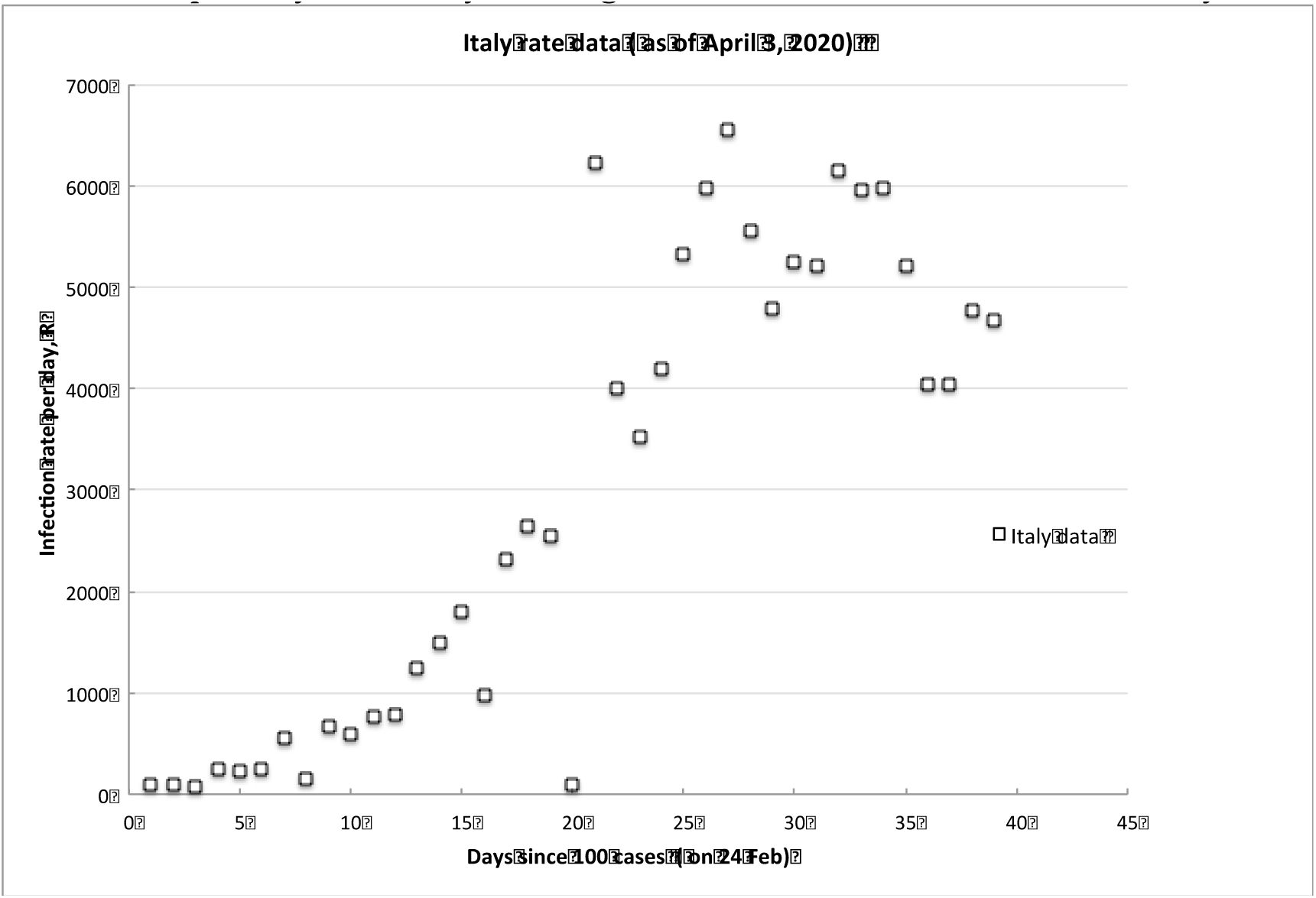
The overall infection rate in Italy

## 5. Comparison to Learning Theory

The results for all countries that show some form of recovery are reported in the Figure 4- graph, which plots as, E*, the non-dimensional infection rate normalized to the initial peak value, versus N* = ε/ ε_T_, the non-dimensional elapsed time of experience/knowledge or risk exposure after the rate has peaked (number of days after peak/day of peak). In relation to the equations (1) and (2) of learning theory above, the infection rate takes the role of the error rate R, the risk exposure time, ε, corresponds to the accumulated experience/knowledge from which we learn and is measured in days, the time of peak ε_T_, is the time for the rate to approach its achievable minimum value, *R*_*m*_ (the lowest or minimum achievable error rate, in equation (2). Based on the available data for China and S Korea, using countermeasures, the overall recovery timescale is about 20-30 days to attain the minimum infection rate of about 50 per day.

**Figure 4.**
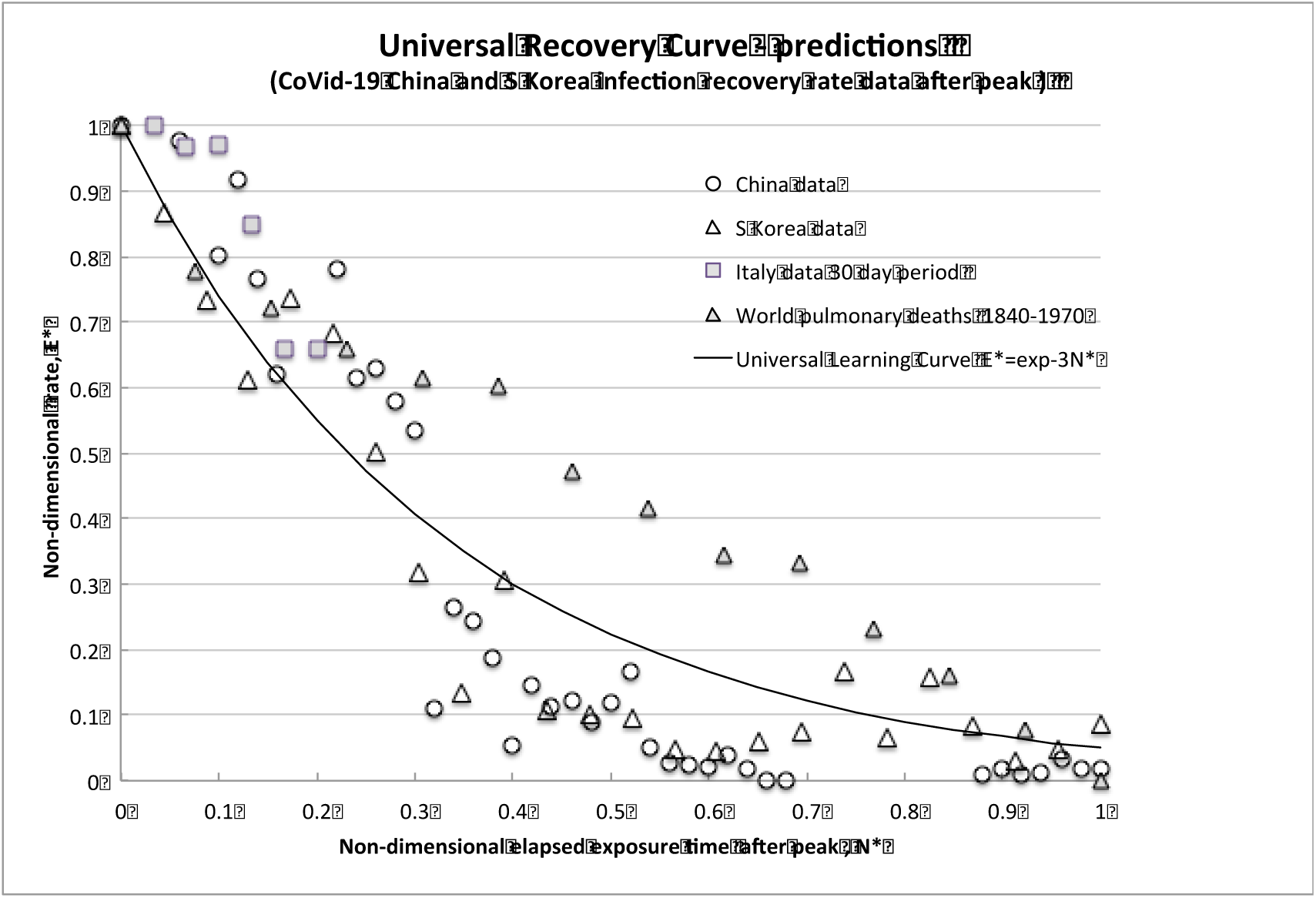
Predicted recovery rate curve (universal learning curve) compared to data.

For further direct comparison, we plot also the reduction curve of the world pulmonary disease death rate per day for 1870-1970 (Source: Human Nature, T.McKeown, April, 1978 as given in Horwitz and Ferleger “Statistics for Social Change”). We can simply think of this overall World data over the years, and its reduction trend, as resulting from many pandemics and multiple outbreaks of influenzas and differing virus strains, that have been more and more successfully treated as we have learned to better control/reduce infections and improved effect recovery, thus steadily reducing the rate.

Despite the huge differences in timescales, the “recovery rate curve” is simply the exponential Universal Learning Curve of equation (3) above, given by E*= exp- KN.

The CoVid-19 pandemic and pulmonary disease recovery rate data all fit with the Universal Learning Curve trend (which is known to fit millions of events with learning). The infection rate, E*, normalized to the initial peak value, as a function of the time elapsed from the peak, N*, normalized to the time of peak are also plotted for China (data circles) and S. Korea (open triangles), and for Italy (squares) based on about a 30–day timescale.

The key point is that all the data follow almost exactly the same decreasing trajectory and, furthermore, the learning curve is nearly the same (K∼3), as previously found for any learning experience on outcomes, accidents, events of any other modern technological system operated by humans. Indeed, the results surprisingly follow the curve developed some ten years after it was first discovered, while working on completely different data.

We can claim that this trend decline due to learning is direct evidence of learning about risk *reduction* also in this case of the pandemic, and call it the Universal Recovery Curve.

To further confirm the URC general theoretical correlation, we next compare to the latest projections for medical resource loads made by complex computer modeling of infections and deaths in the USA (IHME, 2020). As a reasonable surrogate measure, the number of required hospital beds was assumed to be proportional to the number of infections, which daily values were directly transcribed from the website graph (available at covid19.healthdata.org/united-states-of-america). The interval available is a projection from a peak resource use on April 15^th^ out to July 1^st^, 2020, so to be consistent with the actual available country data. The infection rate per day, R, was calculated until attaining an assumed but realistic minimum rate, R_m_, of 50 per day on 10^th^ June (55 days later).

The comparisons of the *observed* decreasing infection rate for both (independent) countries (China data circles, S. Korea open triangles) and the widely used IHME model projections with the Universal Learning Curve shown in the Figures 4 and 5 are compelling. The data fits with learning curve theory, which we know already incidentally fits millions of events, accidents and trends. China, Italy and S. Korea have indeed learned how to control the spread of a viral pandemic. All other countries/systems/people have to do to predict the infection rate evolution is to follow the same trend after first reaching their rate peak. This type of analysis allows countries and systems to compare the effectiveness of their countermeasures implemented to control the pandemics and the related timescales.

**Figure 5.**
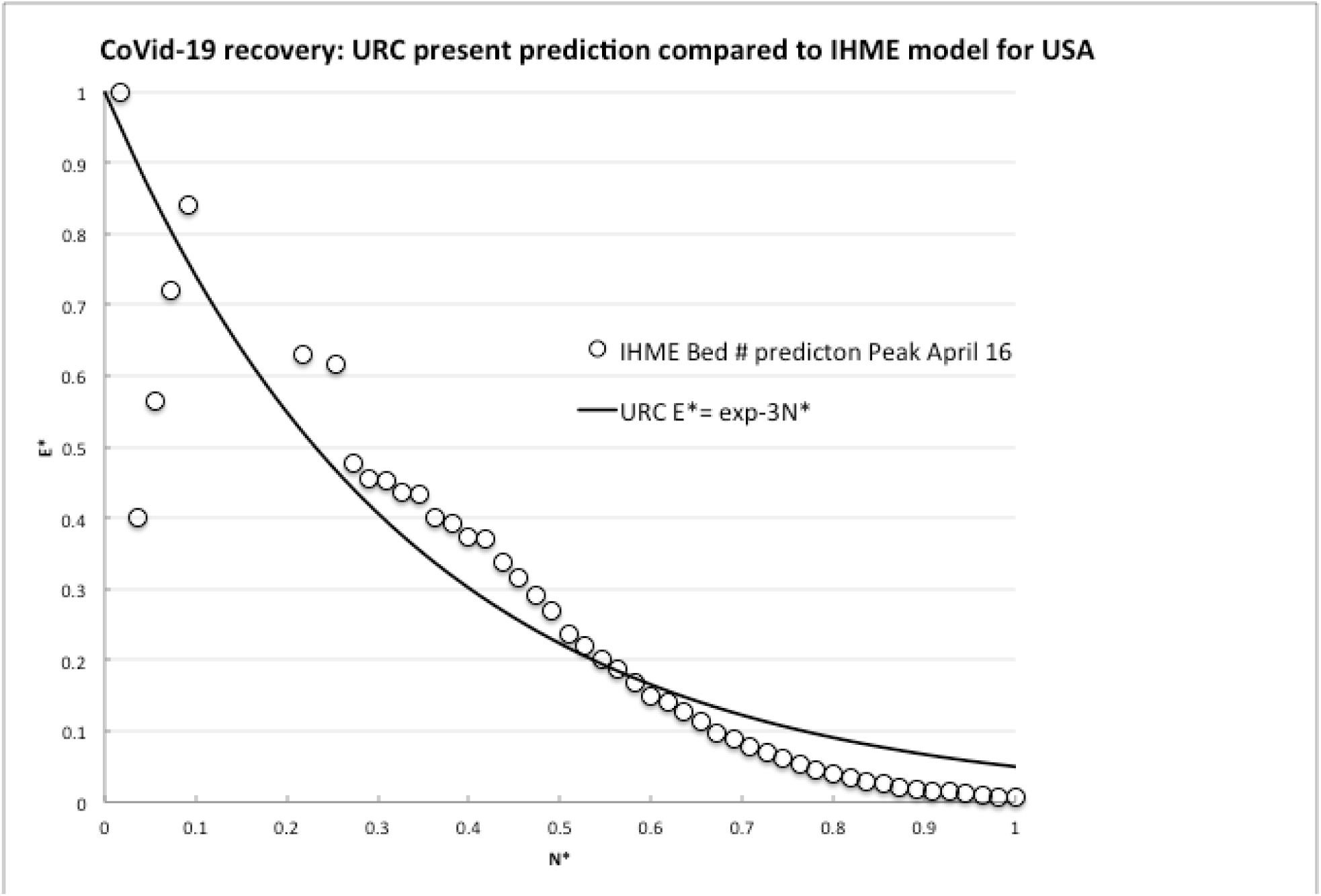
Comparison of learning theory to model predictions of required hospital beds

A word of caution is necessary, however, these numbers cannot be exact and are not meant to be exact. These are just calculated risk estimates, which are subject to uncertainty related to all the many endogenous factors related to the virus spreading, and the actuation and respect of the measures implemented. The numbers provide guidance to thinking about the absolute risk and the best approach to take given the risk is constant unless we do nothing to reduce it!

The strong message here is that the rational and logical approach to dealing with the risk of the occurring pandemic (as with any other risk, for that matter) is to limit own personal and potential exposure, and to minimize both the size and scale of the potentially exposed population. This is precisely what governments and contagious disease experts have been saying all along- but is also what any individual should be doing anyway while exposed to the risks of “normal” life. A sort of ethics of resilience (B. Rajaonah and E. Zio, 2020).

## 6. Recovery trajectory and timescale predictions for Italy

In the above graph of Figure 4, the infection reduction/recovery rate for Italy was estimated to occur in about 30 days, by assuming it was a timeframe similar to that of China. The present prediction is, then, that there is at least a 30-day recovery timeframe as the horizon for the infection rate to get down to a minimum achievable level of say 50 per day (as for China); infection rates should be below 10% of the peak (i.e. about 600 per day) in about 3 weeks from the peak. But this prediction can be effectively monitored and should be updated as more data arises.

In fact, at present, only very few days have elapsed since the seemingly reached peak rate, so it is useful to look at a future trajectory. This can be done as in Figure 6, by plotting the data simply changing the denominator of the non-dimensional elapsed time after the peak rate N* from the 30 days (27 March to 27 April) assumed in analogy to China, to say 20 days (27 March to 6 April) or 40 days (27 March to 6 May) for estimated reduction to the (non-zero) minimum (i.e. 50-100 per day). This sensitivity analysis allows to verify, based on objective data, whether or not any change or improvement in countermeasures is warranted, and by how much. The calculations can assume the infection rate peaked either on about 24 March or March 29, the latter being shown as giving the fewest data points (see Figure 6 above)

**Figure 6.**
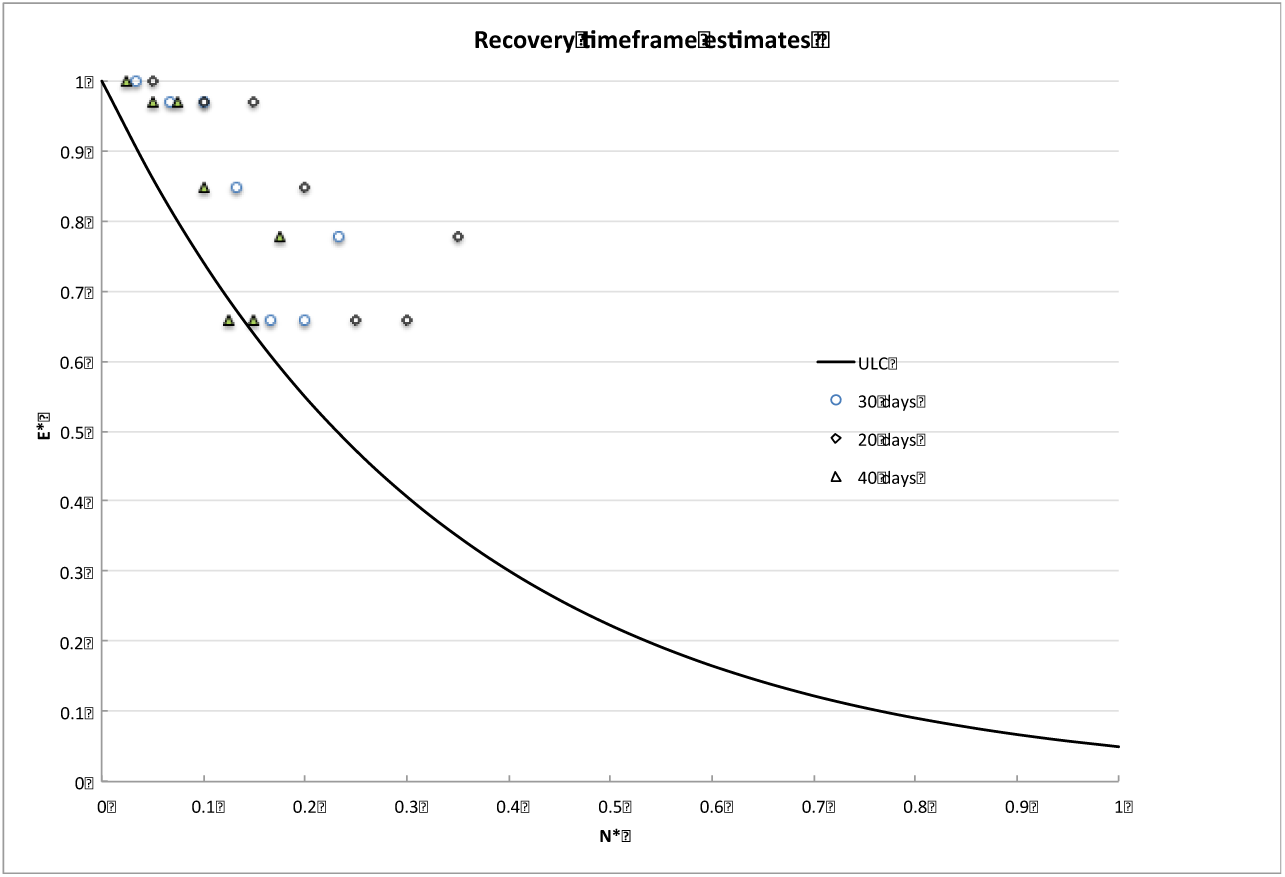
Preliminary calculated recovery trajectories for Italy, and the expected trends based on different peak rate time values ε T of 20,30 and 40 days (data to April 2)

## 7. Conclusions

In this paper, we have originally proposed to adapt Learning Theory for describing the reduction of pandemic infections like that of CoVid-19. A key point is to look at infection rate, as a measure of error outcome, and time, as a measure of experience/knowledge or risk exposure which allows learning. The analyses of the currently available data show that the CoVid-19 infection rate data follow, after peaking, almost exactly the Universal Learning Curve describing the decreasing trajectory of many other instances where humans learn to apply effective countermeasures. More specifically, the learning curve is nearly the same (with universal constant K∼3) as for any learning experience reducing outcomes, accidents and events for other modern technological system operated by humans.

We claim that this trend decline due to learning is direct evidence of learning about risk reduction, also in this case of the pandemic, and call it the Universal Recovery Curve. It can be used to predict the expected time at which the pandemic will be under control, in terms of minimum achievable infection rate, and to test and demonstrate the relative effectiveness of the adopted countermeasures. As such, it is a fundamental tool for risk handling during the development of a pandemic.

## Data Availability

All data are all publicly available, e.g. at Johns Hopkins University website coronavirus.jhu.edu/map

https://www.arcgis.com/apps/opsdashboard/index.html#/bda7594740fd40299423467b48e9ecf6

## Acknowledgement

The authors thank Professor Francesco D’Auria for his many comments and suggestions on this work.

## Footnotes

1 To be clear on terminology: Pandemic-disease affecting the whole world; Epidemic-disease affecting whole communities; Pestilence-a fatal epidemic disease.

2 It turns out to be similar to the assumptions made in the elegant simulations shown at www.washingtonpost.com/graphics/2020/world/corona-simulator/

3 For the mathematically inclined, this function is available in the Excel program in Microsoft Office under the heading HYPGEOMDIST, and is discussed extensively in Edwin Jaynes’ book (see Bibliography)

4 Numerically, the peak corresponds to the complement of the probability of not having more infections (1/e=0.366)

5 For similar pessimistic estimates see, for example, www.kevinmd.com/blog/2020/03/a-covid-19-coronavirus-update-from-concerned-physicians.

6 For similar realistic estimates see, for example, https://wmbriggs.com/post/29830/

## Notes

### Competing Interest Statement

The authors have declared no competing interest.

### Funding Statement

No external funding

